# Modern Contraceptive Utilization and Associated Factors Among Reproductive-age Women In Ethiopia, Evidence from PMA Ethiopia 2023 CS survey

**DOI:** 10.64898/2025.11.30.25341252

**Authors:** Aliyu DessalewAli, Solomon Abrha Damtew, Mihretu Alemayehu

## Abstract

**Background:** Unmet contraceptive needs continue to be a serious public health concern in the world, despite modern contraception’s critical role in enhancing maternal and child health. Existing publications focus on out-of-date data and overlook community-level factors, such as community FP knowledge. Therefore, this study employs the most recent Performance Monitoring for Action (PMA) Ethiopia 2023 survey to assess both individual and community-level variables of modern contraception use.

**Methods:** The study used nationally representative PMA 2023 cross-sectional data with a two-stage cluster sampling technique. Data were cleaned and prepared for analysis using STATA version 14. 5,250 reproductive-age, sexually active, and non-pregnant women were included in the study. Descriptive statistics and bivariate analysis were performed. To identify associated factors, multilevel binary logistic regression analysis was used. ICC, MOR, PVC, log-likelihood, and deviance were used for model comparison and fitness. To identify significantly associated factors with modern contraceptive utilization, AOR with 95% CI was used. A *p*-value of 0.05 was used to declare significance.

**Results:** The magnitude of modern contraceptive utilization in Ethiopia was 41.7%. The multilevel analysis demonstrated that women aged 35-49 years [AOR = 0.50; 95% CI: 0.35-0.71], not currently married [AOR = 0.45; 95% CI: 0.33-0.61], with parity of more than 5 children [AOR = 0.54; 95% CI: 0.35-0.83], and residing in pastoralist regions [AOR = 0.31; 95% CI: 0.19-0.51] were negatively associated with modern contraceptive utilization.

In contrast, household size of 4-5 members [AOR = 1.44; 95% CI: 1.15-1.81], mixed feelings if pregnant now [AOR = 2.56; 95% CI: 1.98-3.31], unhappy feelings if pregnant now [AOR = 2.13; 95% CI: 1.74-2.61], recent health facility visit [AOR = 1.53; 95% CI: 1.19-1.97], moderate family planning knowledge [AOR = 1.35; 95% CI: 1.01-1.81], Muslim religion [AOR = 1.48; 95% CI: 1.13-1.96], residing in communities with high levels of women’s education [AOR = 1.82; 95% CI: 1.28-2.60], and communities with high media exposure [AOR = 1.42; 95% CI: 1.00-2.00] were positively associated with modern contraceptive utilization.

**Conclusions:** In Ethiopia, modern contraceptive utilization still remains below national target, influenced by both individual and community-level factors. Individual-level factors were women’s age, married status, religion, parity, visited health facility, household member size, reaction if got pregnant and FP knowledge. Community-level factors such as region, community women education, and community media exposure were identified. These findings highlight the significance of multilevel, context-specific interventions. Older women, women with high parity, and those who live in pastoral areas require special care.

## Background

Modern contraceptive methods are technological advances developed to allow couples to engage in sexual activity according to their natural impulses and desires while reducing the risk of unintended pregnancy(1). Modern contraceptive methods, such as oral pills, implants, injectables, condoms, and intrauterine devices (IUDs), are proven to be effective in preventing pregnancy and are linked with improvements in maternal and child health outcomes(2,3). The use of modern contraceptive methods is critical for improving reproductive health outcomes, preventing unwanted pregnancies & associated risks, and empowering women to make educated reproductive decisions(4). Modern contraceptive methods are also fundamental for enhancing maternal and child health outcomes, advancing gender equality, and achieving global health goals(5).

Globally, 966 million women use contraceptives, the number of women using modern contraception nearly doubled from 467 million (35%) in 1990 to 874 million (45%) in 2019. However, 164 million women desire to delay or avoid conception and are not taking any kind of contraception, indicating an unmet demand for family planning(6).

Between 1990 and 2021, the proportion of family planning needs met by modern methods increased globally from 67% to 77.5%, although significant gaps remain, particularly in low-income regions like sub-Saharan Africa, where only 58% of needs are met (2,3). In sub-Saharan Africa, the proportion of women who have their need for family planning satisfied with modern methods (SDG indicator 3.7.1) continues to be among the lowest in the world at 56 per cent. However, 22 of the 41 countries where less than half of women who needs to prevent pregnancy use modern contraceptive methods are in sub-Saharan Africa(2).

A community-based cross-sectional survey conducted in the Ho district of west Ghana found that 89.8% of married women used modern contraceptives(7). A community-based cross-sectional survey of reproductive-age women in Yaoundé, Cameroon revealed that 58.9% utilized modern contraceptive methods(8).

By 2030, Ethiopia intended to achieve sustainable development goal 3.7 which is ensuring universal access to sexual and reproductive health-care services, including for family planning, information and education, and the integration of reproductive health into national strategies and programs. Especially, SDGs 3.7.1 reproductive age (aged 15–49 years) who have their need for family planning satisfied with modern methods(9).

Ethiopia has made notable progress in increasing access to modern contraceptives over the past two decades. The modern contraceptive prevalence rate (mCPR) among married women rose from 6% in 2000 to 41% in 2021(1). However, despite this advancement, the overall use of modern contraceptives remains suboptimal, with significant regional disparities and urban-rural differences(10). A study conducted in Ethiopia using the 2019 EDHS data revealed that modern contraceptive use among Ethiopian women of reproductive age was 28%(11).

Although numerous studies(4,10,15–19) have investigated individual-level determinants of modern contraceptive utilization-such as age, education, marital status, parity, wealth index-less attention has been paid to contexual service-related factors. In particular, variables such as community level women FP knowledge.

Most researchers studied modern contraceptive use among Ethiopian women of reproductive age but most studies either analyze all women or limit their analysis to married women without accounting for their sexual activity in the past year. The groups studied in these research studies do not necessarily include women who are at risk of pregnancy at the moment. The inclusion of sexually inactive women in these studies biases the results by underestimating the real need for contraception. The study focuses on sexually active women during the last year in order to establish an accurate and relevant measurement of modern contraception utilization among individuals who are most likely to become pregnant unintentionally.

Many previous studies examining contraceptive utilization in Ethiopia are outdated or based on earlier rounds of demographic surveys, failing to capture recent changes in family planning programs and sociocultural dynamics (4,16,20–25). Moreover, most research has predominantly focused on individual-level predictors, with limited exploration of community-level influences (23).

The Performance Monitoring for Action (PMA) project offers up-to-date and high-quality data on family planning and reproductive health indicators. Unlike older surveys, PMA uses innovative mobile-based data collection to provide rapid, frequent, and nationally representative estimates (24). However, analyses using the latest PMA Ethiopia 2023 survey data remain scarce, and comprehensive investigations integrating both individual and community-level factors are needed.

Thus, the current study aims to determine the magnitude of modern contraceptive utilization and identify associated factors among reproductive-age women in Ethiopia using nationally representative data from PMA Ethiopia 2023 cross-sectional survey. By addressing individual, and community-level determinants, this research intends to fill important gaps and inform policy and programmatic decisions to enhance modern contraceptive use in Ethiopia.

## Methods and Data Sources

### Study Design, Population and Sample Size

This study utilizes secondary cross-sectional data from the PMA Ethiopia 2023 survey,which is a reliable and recent source of nationally representative survey designed to capture comprehensive data on reproductive, maternal and newborn health indicators in Ethiopia. Data were collected by Performance Monitoring for Action (PMA) project from November 2023 to January 2024. This project implemented in collaboration with Addis Ababa University, Johns Hopkins University, and the Federal Ministry of Health. The funding is provided by the Bill & Melinda Gates Foundation (25). The data are freely available for public on Johns Hopkins University (JHU) Data Repository, which is open-access platform.

The data were collected from reproductive-age, sexually active, and non-pregnant women in Ethiopia from twelve regions: namely: Tigray, Afar, Amhara, Oromia, Somali, Benishangul-Gumuz, SNNP, Gambela, Harari, Addis Ababa, Dire Dawa, Sidama, Southwest Ethiopia, and Central Ethiopia.

First the data set was kept by gender(keep if gender==2) then kept by age(keep if age>=15 & age<=49). By using both variables “last_time_sex” and “last_time_sex_value”, generate a variable “last_sex_days”, then we generate a variable “sex_inactive” which has two categories i.e 1 “sexually inactive” 0 “sexually active”. After that we exclude sexually inactives(drop if sex_inactive==1) and women who never had sex (drop if why_not_usingnosex==1). Next to this, we kept the data by pregnancy (keep if pregnant==0), then we exclude infecund and menopausal women (drop if sex_inactive==1 and drop if why_not_usingnosex==1). Finally, we kept the data by both household questionnaire and female questionnaire (keep if HHQ_result==1 & FRS_result_cc1==1).

After missings are managed,the preliminary analysis and data cleaning resulted a final sample size of 5,250 who were sexually active and non-pregnant women by the time of the PMA Ethiopia(2023) survey.

A representative sample survey Performance Monitoring for Action (PMA-Ethiopia 2023) was used to provide national level reports. All women between the ages of 15 and 49 who reside in the chosen households were included in the PMA-Ethiopia survey. A two-stage stratified cluster sampling method used to select enumeration areas. A complete census was conducted in the selected enumeration areas followed by a selection of 35 households per enumeration area using simple random sampling. All reproductive age women were interviewed after the household survey.

A total of 280 EAs were chosen in the first stage, with independent selection in each sample stratum and a probability proportional to EA size. In the second round of selection, 35 HHs were drawn at random from the newly formed household list using random number generator software. All females aged 15 to 49 who were either long-term residents of the selected HH or guests who slept there the night before the survey were eligible to participate in the interview.

For this study, a sample of 5,333 female individuals between the ages of 15 and 49, sexually active and who are not currently pregnant and complete both household and female questionnaire were suitable for our analysis by considering our purpose were chosen. Those women who were not in the reproductive-age group, sexually inactives, pregnants, infecund and menopausal women, and those women who did not completed both household and female questionnaire were excluded from the analysis. Thus, out of the total 20,535 females included in the PMA 2023, 11,271 females who were not in the reproductive-age group were dropped at initial step. Following the exclusion of 3,163 sexually inactives and those women who had never sex, 765 females who were pregnant at the time of the survey, 2 infecund and menopausal women and 1 females who did not completed the questionnaire were also dropped, leaving an unweighted sample size of 5,333 women. We used sampling weight to address the disproportionate sample allocation in the PMA, resulting in a final weighted sample size for this study of 5,250(Figure 1).

**Figure 1:**
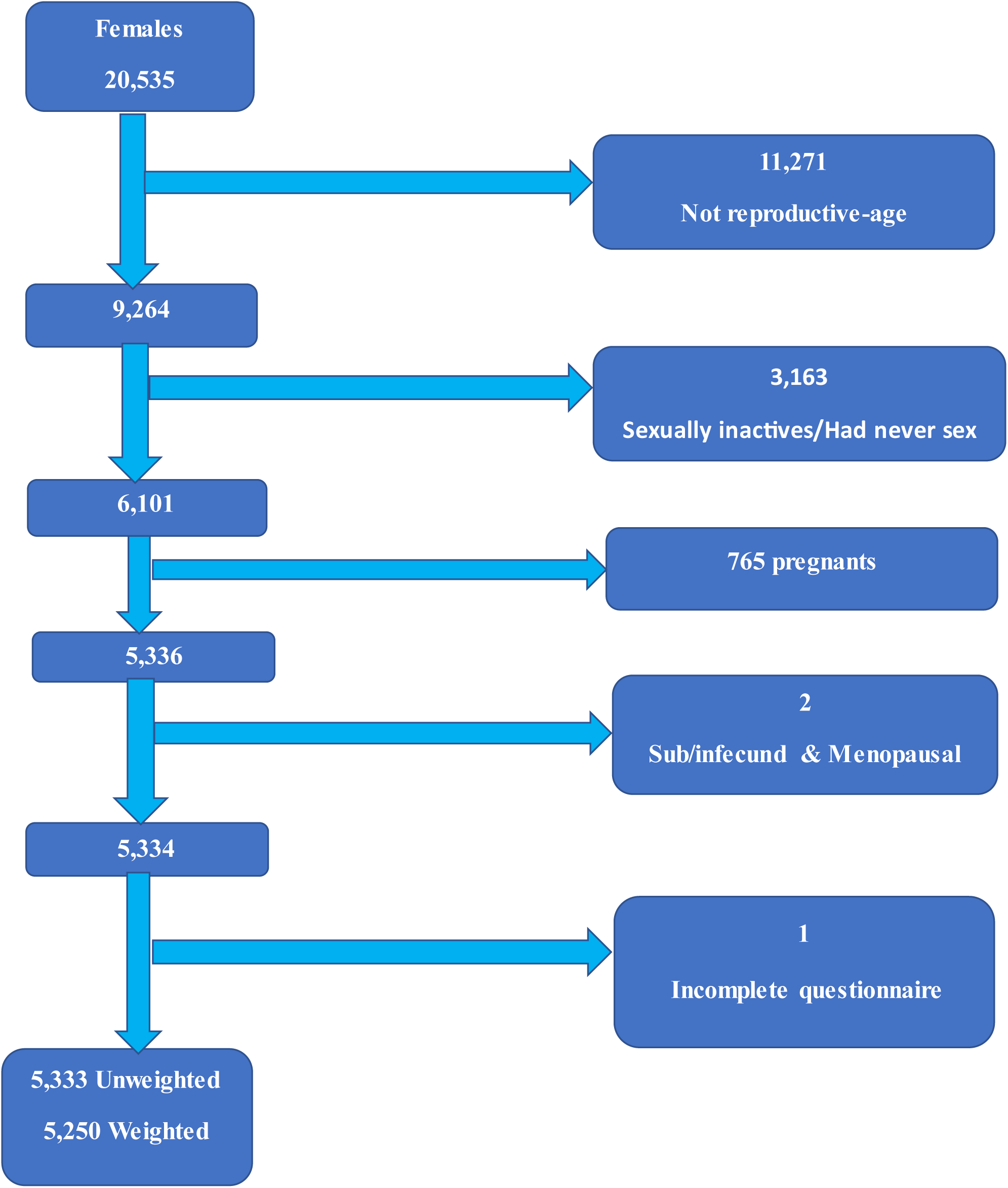
S**c**hematic **presentation of the women included in this study using the 2023 PMA data**

**Figure 2:**
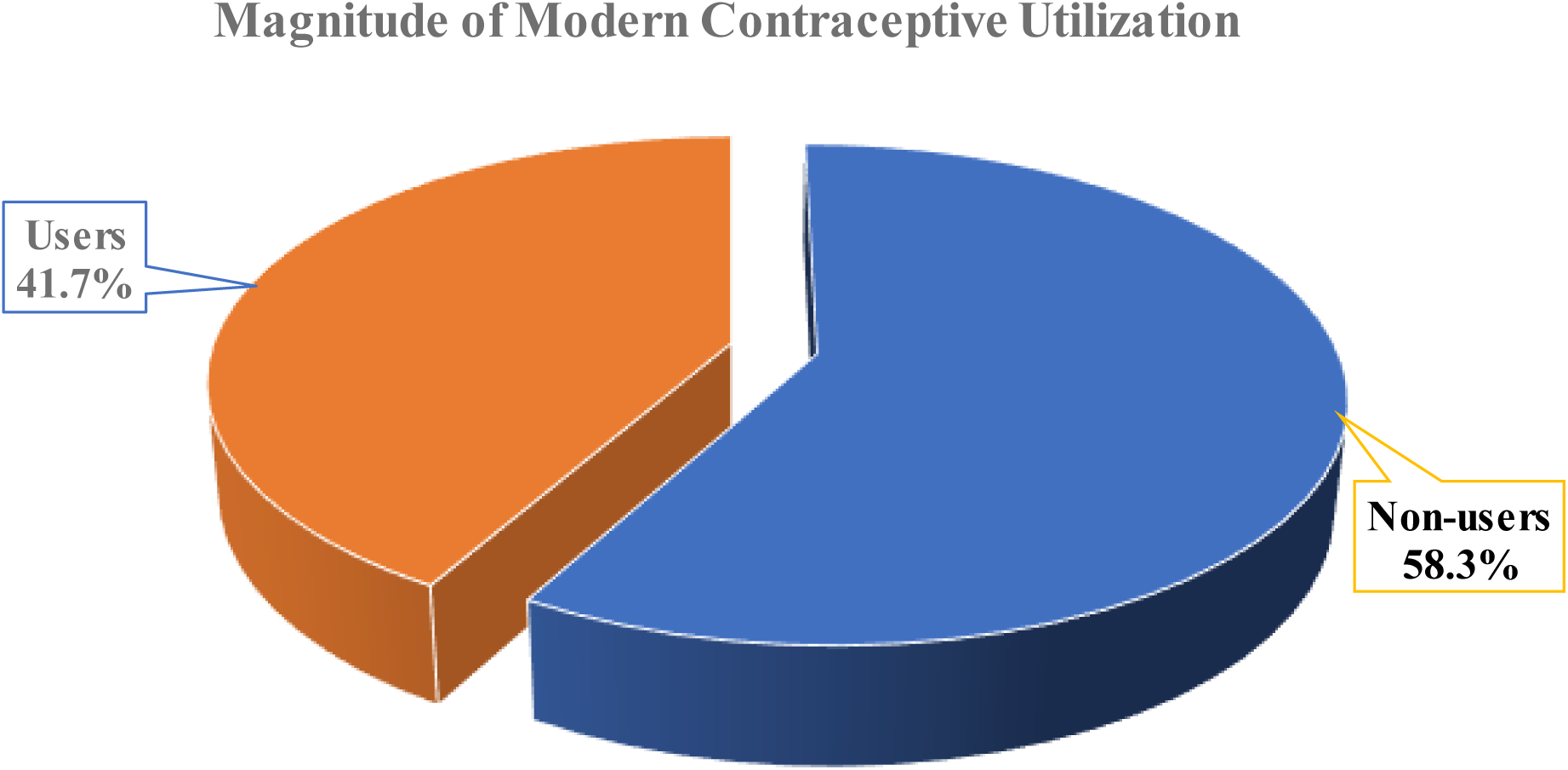
Proportion of magnitude of modern contraceptive utilization among reproductive-age women in Ethiopia,2025(n=5,250)

## Variables

### Outcome variable

The main outcome variable of this study was modern contraceptive utilization (users and non-users). In this study, a woman who had been utilizing any of modern contraceptive methods such as female sterilization, male sterilization, IUCD, injectable, implants, pills, male condom, female condom, emergency contraception, and standard days method in the past 12 months during PMA Ethiopia CS 2023 survey, considered as “modern contraceptive users”. Modern contraceptive users were coded as 1 “Yes”, non-users were coded as 0 “No”.

### Independent Variables

Independent variables were classified into individual-level variables and community-level variables broadly. Individual-level independent variables further categorized into socio-demographic characteristics variables, fertility and SRH characteristics variables, and contraceptive use characteristics variables.

❖ Socio-demographic characteristics
  - Women’s age, Marital status, Women’s education level, Religion, Wealth quintile, and Household member size.
❖ Fertility and SRH characteristics
  - Parity, Reaction if got pregnant, Age at first FP use, Age at first sex, Visited facility
  - Parity was generated by recoding a variable “birth events”.
❖ Contraceptive use related characteristics
  - FP knowledge and Media exposure
  - FP knowledge and Media exposure were created composite variables.

**Community-level (aggregate) factors**

- Region, Residence, Community women education, Community media exposure, Community FP knowledge, Community wealth index
- The aggregate community-level explanatory variables (community women’s education, community partner’s education, community wealth index, community FP knowledge and community media exposure) were constructed by aggregating individual-level characteristics at the community (cluster) level.

### Analysis and Measurement

Data was processed using STATA version 14 software to ensure its accuracy and consistency. The PMA Ethiopia CS 2023 dataset was initially cleaned to remove any missing or inconsistent values of key variables. Furthermore, variables were recoded to provide biologically plausible categories. This was followed by chi-square test checking appropriate categories were merged to make cell sample size adequacy.

Descriptive statistics were used to describe the study population. Frequencies and percentages were computed to characterize the study population. Chi-square test statistics and cross-tabulation were computed to see the overall association/relationship of the independent variables with the dependent variable (modern contraceptive utilization).

At bivariate analysis a *p-*value cut of 0.25 was used to select candidate variable for multilevel multivariable logistics regression analysis. Multi-collinearity was checked by using variance inflation factor (VIF<10).

Multilevel binary logistics regression was used to identify significantly associated factors of modern contraceptive utilization. Multilevel logistic regression model in a combination of both fixed effect (a measure of association) and random effect (a measure of variation) were performed. EA’s was treated as random effects. The random effects were measured by the intra-class correlation coefficient (ICC), median odds ratio (MOR), and proportional change in variance (PVC).

Four models were fitted. Model 0 (null model), with no determinants (random intercept) to estimate random variation in the intercept and ICC. Model 1 – only included individual-level variables, Model 2 – only included community-level variables to estimate the community-level characteristics, and finally, Model 3 – included both individual-level and community-level variables adjusted for both.

The information criteria’s Akaike Information Criteria (AIC) and Schwarz’s Bayesian Information Criteria (BIC) were used to compare the models to choose the best fitted. The best-fit model was the model with the lowest AIC/BIC and highest loglikelihood. The best-fit model was also compared by deviance, and the best fit was a model with low deviance. Model fitness were checked by Hosmer and Lemeshow goodness of fit test. Results were presented in the form of percentage, and odds ratio with 95% CI. Significance was declared at a significance level of 0.05.

### Data Quality Assurance

During PMA Ethiopia 2023 survey, data were collected by well experienced field staff and resident enumerators using smartphones equipped with a customized Open Data Kit (ODK) platform known as PMA Collect, which enables real-time data generation and timely feedback. Standardized and pre-tested questionnaires were administered in three local languages: Amharic, Afan Oromo, and Tigrigna. To ensure data quality, field supervisors conducted regular oversight, including weekly error progress reports, prompt responses to issues, close monitoring during the listing and data collection phases for household and female questionnaires, as well as 10% re-interviews and random spot checks.

## Result

### Socio-demographic characteristics of the respondents

From PMA Ethiopia 2023 dataset, a total of 5,250 reproductive-age women are included in this study. Respondents were 31.45 years old on average, with an SD of 8.14 years. Of those, the majority (64.8%) of respondents were lived in rural areas. Regionally, Oromia (17.83%) had the largest percentage of respondents. In terms of age, 41.14% of the women were between the ages of 25 and 34 years. The majority of respondents were identified as Christians (69.39%), followed by Muslims (29.37%). The majority of the respondents (90.19%) were current in union (married and living with partner). The educational status showed that 37.31% attained primary education. Regarding wealth status, 23.68% were in the highest wealth quintile, while 21.35% were in the lowest. (See Table 1 below)

**Table 1:** Socio-demographic characteristics of reproductive age women in Ethiopia, PMA 2023 Ethiopia (weighted, n = 5250)

| Variables | Category | Frequency | Percent |
| --- | --- | --- | --- |
| Age | 15 to 24 years | 1,113 | 21.20 |
|  | 25 to 34 years | 2,160 | 41.14 |
|  | 35 to 49 years | 1,977 | 37.66 |
| Residence | Urban | 1,848 | 35.2 |
|  | Rural | 3,402 | 64.8 |
| Region | Tigray | 451 | 8.59 |
|  | Afar | 238 | 4.53 |
|  | Amhara | 834 | 15.89 |
|  | Oromia | 936 | 17.83 |
|  | Somali | 335 | 6.38 |
|  | Benishangul-Gumuz | 95 | 1.81 |
|  | SNNP | 568 | 10.82 |
|  | Gambela | 369 | 7.03 |
|  | Harari | 53 | 1.01 |
|  | Addis | 428 | 8.15 |
|  | Dire Dawa | 75 | 1.43 |
|  | Sidama | 325 | 6.19 |
|  | Southwest Ethiopia<br>People | 396 | 7.54 |
|  | Central Ethiopia | 147 | 2.80 |
| Religion | Christians | 3,643 | 69.39 |
|  | Muslims | 1,542 | 29.37 |
|  | Other traditional | 45 | 0.86 |
|  | No religion | 20 | 0.38 |
| Marital status | Currently in union | 4,735 | 90.19 |
|  | Previously married | 257 | 4.90 |
|  | Never married | 258 | 4.91 |
| Education attained | No Education | 1,917 | 36.51 |
|  | Primary | 1,959 | 37.31 |
|  | SecondaryPlus | 1,374 | 26.17 |
| Household member size | 1 to 3 members | 1,273 | 24.25 |
|  | 4 to 5 members | 1,970 | 37.52 |
|  | >= 6 members | 2,007 | 38.23 |
| Wealth quintile | Lowest quintile | 1,121 | 21.35 |
|  | Lower quintile | 974 | 18.55 |
|  | Middle quintile | 972 | 18.51 |
|  | Higher quintile | 940 | 17.90 |
|  | Highest quintile | 1,243 | 23.68 |

### Magnitude of Modern Contraceptive Utilization

Out of 5250 participants,2188(41.7%) of reproductive-age women (15–49 years) in Ethiopia are currently using modern contraceptive methods, while the remaining 3062(58.3%) are not.

### Fertility and SRH Characteristics

Regarding parity, 47.07% of respondents had 0–2 children. Most women had their first sexual intercourse between 15–19 years (61.45%), and a substantial proportion started using family planning (FP) after age 30 (41.15%). Regarding reactions towards pregnancy, 45.12% of the respondents reported they would feel happy if they became pregnant now. Facility visits for FP services were reported by 63.03% of respondents.(See Table 2 below)

**Table 2:** Fertility and SRH Characteristics of reproductive-age women in Ethiopia.

| Variables | Category | Frequency | Percent |
| --- | --- | --- | --- |
| Parity | 0 to 2 children | 2,472 | 47.09 |
|  | 3 and 5 children | 1,768 | 33.68 |
|  | > 5 Children | 1,010 | 19.24 |
| Age at first sex | <=14 years | 581 | 11.07 |
|  | 15-19 years | 3,226 | 61.45 |
|  | 20-24 years | 1,085 | 20.67 |
|  | >=25 years | 358 | 6.82 |
| Age at first FP use | <=14 years | 45 | 0.86 |
|  | 15-19 years | 1,226 | 23.35 |
|  | 20-29 years | 1,871 | 35.64 |
|  | >=30 years | 2,108 | 40.15 |
| Reaction if pregnant now | Happy | 2,369 | 45.12 |
|  | Mixed | 610 | 11.62 |
|  | Unhappy | 2,271 | 43.26 |
| Visited facility | No | 1,941 | 36.97 |
|  | Yes | 3,309 | 63.03 |

### Contraceptive use Characteristics

Regarding family planning knowledge, 62.02% of the respondents had moderate knowledge, 23.03% had poor knowledge, and 14.95% had good knowledge.

Majority of the respondents(66.25%) had no media exposure, while 33.75% of respondents reported they had exposure to family planning messages via at least one media channel. (See Table 3 below)

**Table 3:**
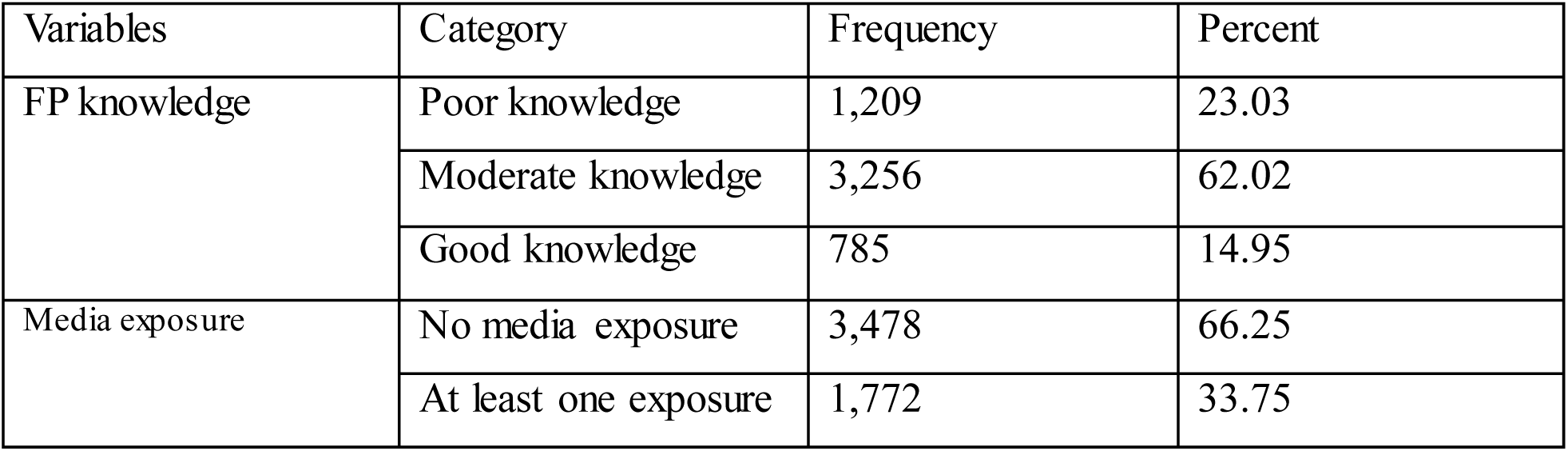
Contraceptive use Characteristics of reproductive-age women in Ethiopia, PMA Ethiopia 2023.

| Variables | Category | Frequency | Percent |
| --- | --- | --- | --- |
| FP knowledge | Poor knowledge | 1,209 | 23.03 |
|  | Moderate knowledge | 3,256 | 62.02 |
|  | Good knowledge | 785 | 14.95 |
| Media exposure | No media exposure | 3,478 | 66.25 |
|  | At least one exposure | 1,772 | 33.75 |

### Community-level Characteristics

Enumeration areas were taken as a unit for analysis. Among 280 EAs, 52.19% of the cluster had high community-level women’s education. Half (50.67%) of the cluster were classified as high poverty. Half (50.17%) of the cluster had high community-level family planning knowledge. Of the total 280 EAs, 50.48% of the cluster had high community-level media exposure. (See Table 4 below)

**Table 4:**
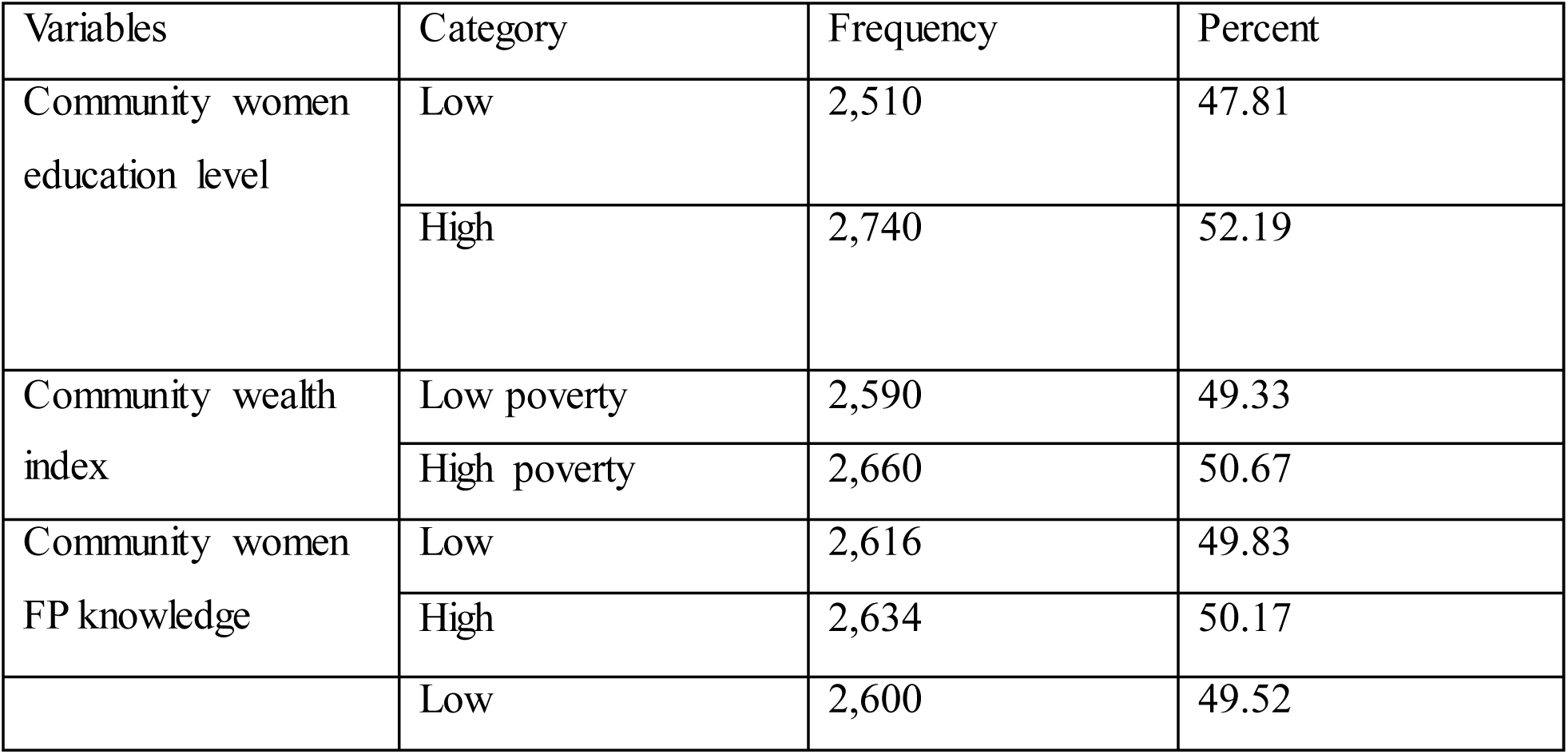
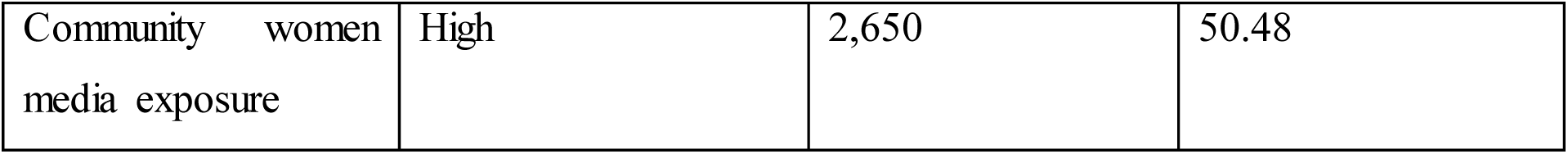
Descriptive summary of Community-level aggregate Characteristics among reproductive age women, PMA Ethiopia 2023.

### Factors associated with Modern contraceptive utilization

In the bivariate logistic analysis, several individual-level and community-level factors were found to be significantly associated with modern contraceptive utilization at a p-value less than 0.25 (p< 0.25). These included women’s age, educational status, wealth quintile, region, residence, parity, marital status, religion, household member size, age at first sex, age at first FP use, visited facility, reaction if got pregnant now, FP knowledge, and media exposure. Additionally, all examined community-level factors; community FP knowledge, community wealth index, community women’s education, and community media exposure—showed significant associations with modern contraceptive utilization. These variables were selected as candidate variables for further analysis in the multilevel binary logistic regression model. The table 5 below shows the crude odds ratio (COR), 95% confidence intervals, and p-values.

**Table 5:**
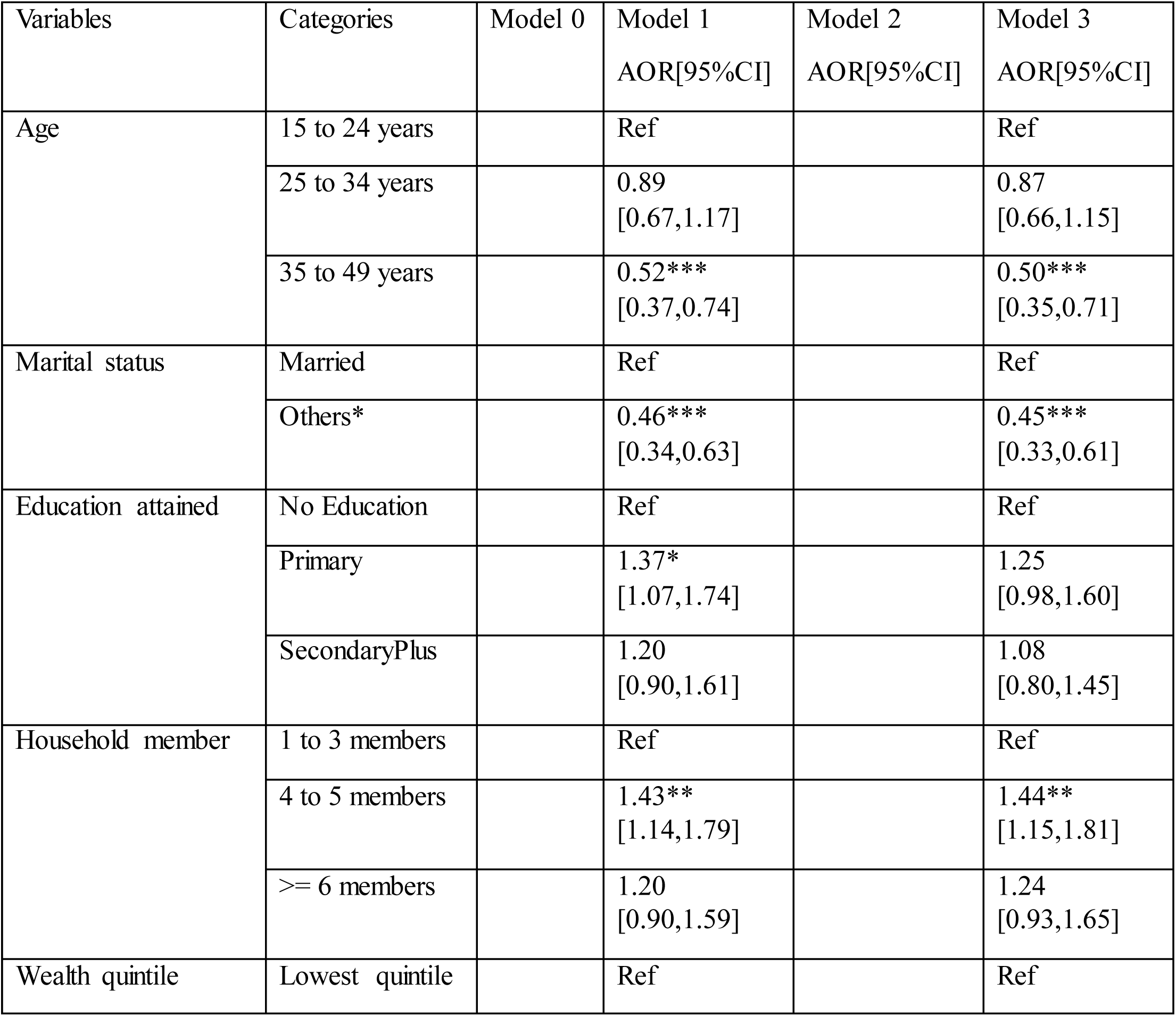
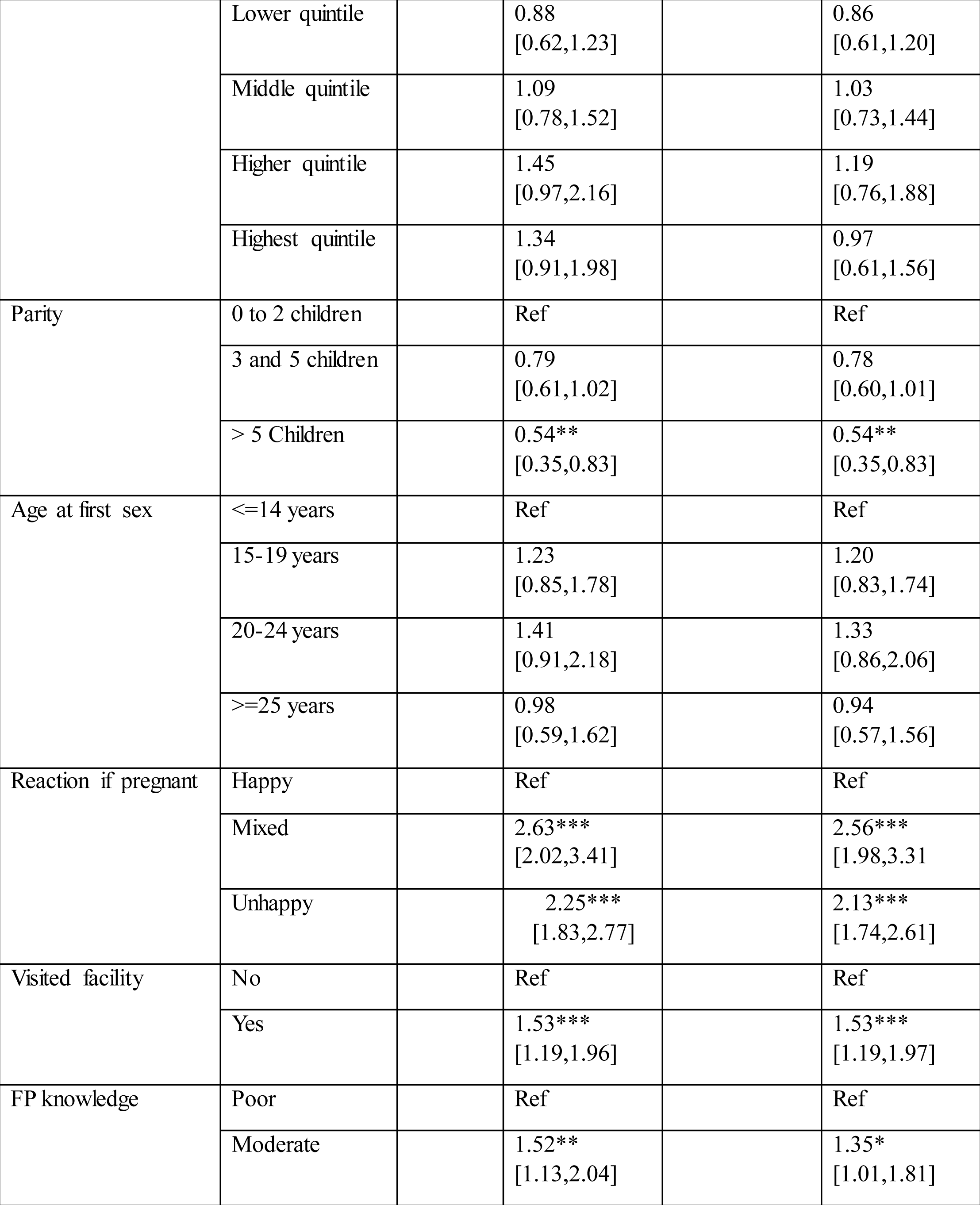
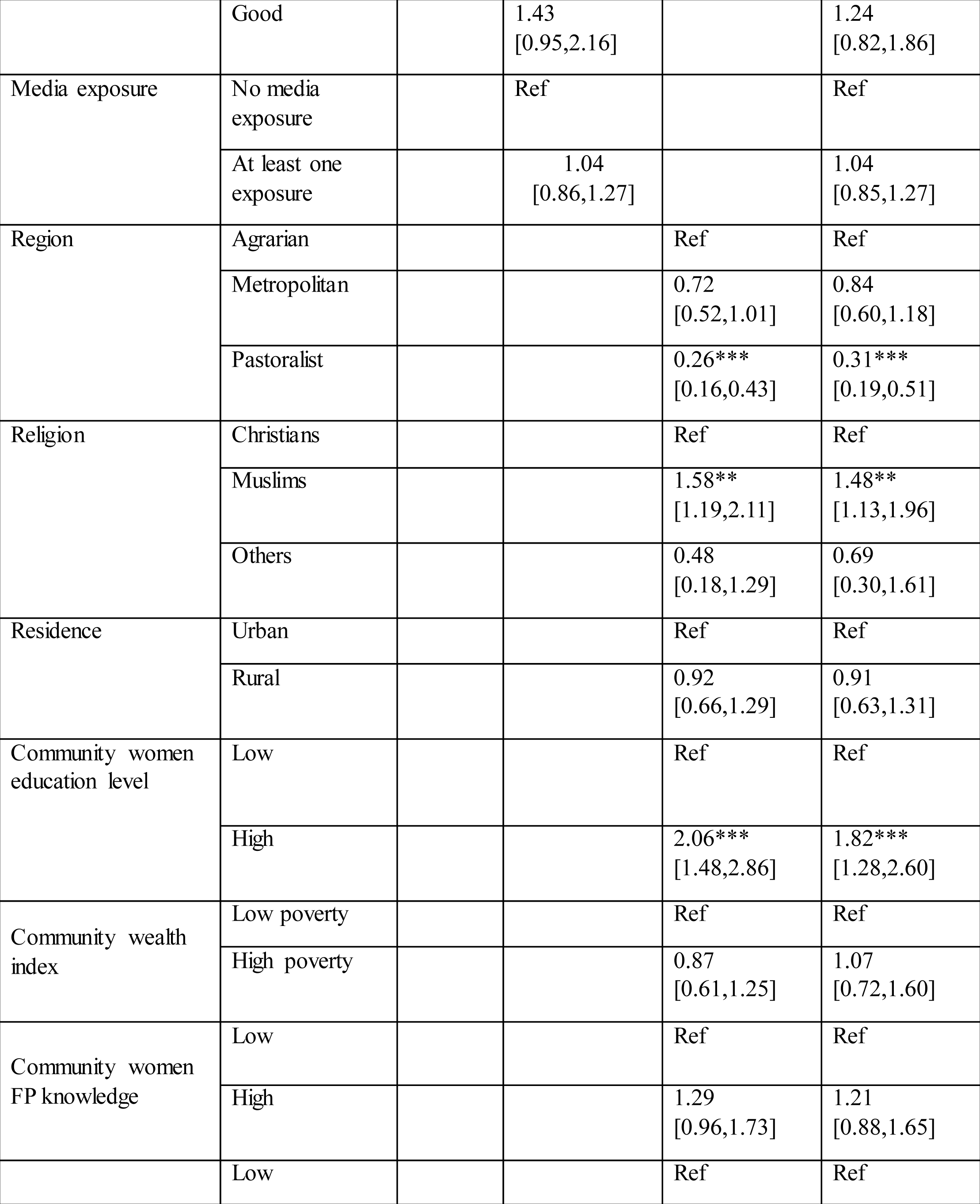
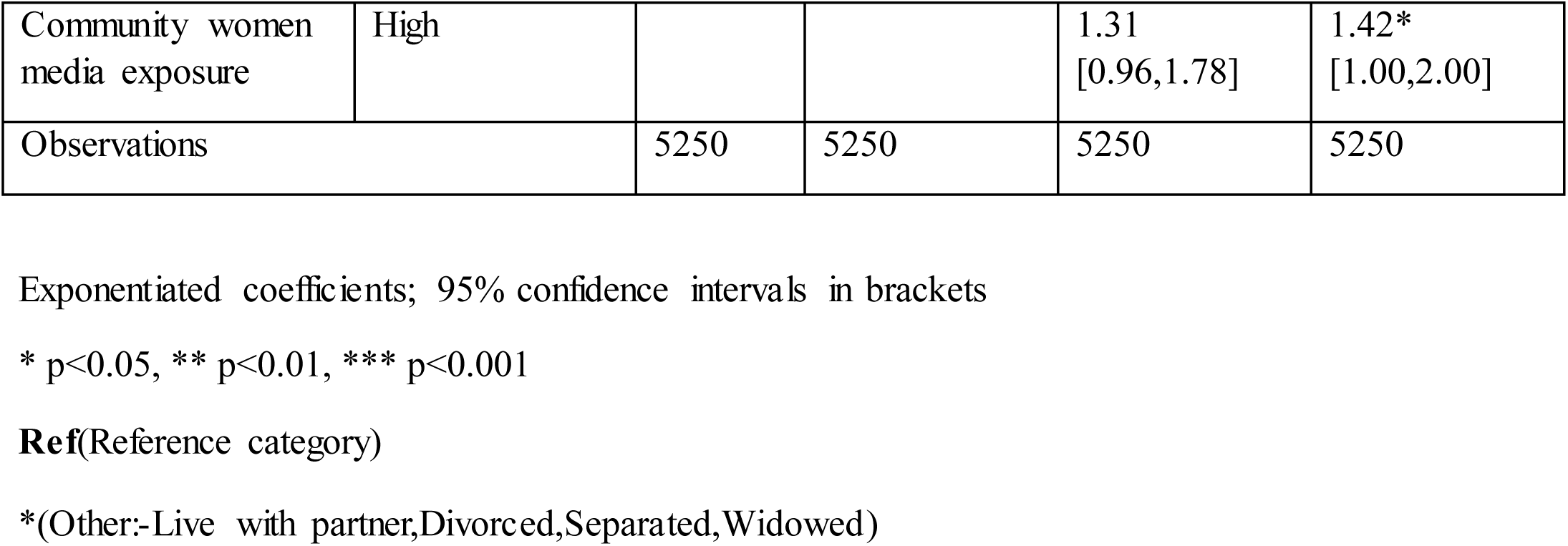
Multilevel Binary Logistics Regression Result for factors affecting modern contraceptive utilization among reproductive-age women, Evidence from CS 2023 data.

### Multilevel binary logistic regression analysis results

A total of 5,250 women were included in the final multilevel binary logistic regression analysis. Four hierarchical models were fitted: a null model(Model 0), an individual-level model(Model 1), a community-level model(Model 2), and a full model including both individual and community-level variables(Model 3). Model 3 being the final adjusted model, accounting for both individual– and community-level variables.

### Fixed-effect Analysis Result

#### Individual-Level Factors

- **Age:** women age 35 to 49 years were 50% less likely to use modern contraceptives as compared to those women aged 15 to 24 years (AOR = 0.50, 95% CI:0.35–0.71, p<0.001).
- **Marital status:** women who were not married had 55% lower odds of utilizing modern contraceptives as compared to those married women (AOR = 0.45, 95% CI: 0.33–0.61, *p<0.001)*.
- **Religion:** muslim women had 48% increased odds of utilizing modern contraceptives compared to Christians women (AOR = 1.48, 95% CI: 1.13–1.96, p<0.01).
- **Household member size:** women living in a household with 4 to 5 members had 44% higher odds of utilizing modern contraceptives (AOR = 1.44, 95% CI: 1.15–1.81, p<0.01) as compared to those women living in 1 to 3 household member.
- **Parity:** women with more than 5 children were 46% less likely to use modern contraceptives (AOR = 0.54, 95% CI: 0.35–0.83, p<0.01) as compared to those with 0-2 children.
- **Reaction if pregnant now**
  ✓ Women who felt mixed (happy and unhappy) about becoming pregnant had 2.56 times more odds of using modern contraceptives (AOR = 2.56, 95% CI: 1.98–3.31, p<0.001) as compared to those who would be happy.
  ✓ Women who would be unhappy if pregnant had 2.13 times more odds of using modern contraceptives (AOR = 2.13, 95% CI: 1.74–2.61, p<0.001) as compared to those women feel happy.
- **Visited facility:** women who visited health facility had 53% higher odds of utilizing modern contraceptives (AOR = 1.53, 95% CI: 1.19–1.97, p<0.001) as compared to those women who did not.
- **FP knowledge:** women with moderate FP knowledge had 35% higher odds of using modern contraceptives (AOR = 1.35, 95% CI: 1.01–1.81, p<0.05) as compared to those women with poor knowledge.

#### Community-Level Factors

- **Region:** women in the pastoralist regions were 69% less likely to use modern contraceptives (AOR = 0.31, 95% CI: 0.19–0.51, p<0.001) as compared to those women in the agrarian regions.
- **Community Women’s Education Level**: Women who resides in communities with high level of women’s education had 82% increased odds of using modern contraceptives (AOR = 1.82, 95% CI: 1.28–2.60, p<0.001) as compared to those women in low education communities.
- **Community Women’s Media Exposure:** Women who reside in communities with high media exposure had 42% increased odds of using modern contraceptives (AOR = 1.42, 95% CI: 1.00–2.00, *p<0.05)* as compared to those women in low media exposure communities. (See Table 6 below)

**Table 6:**
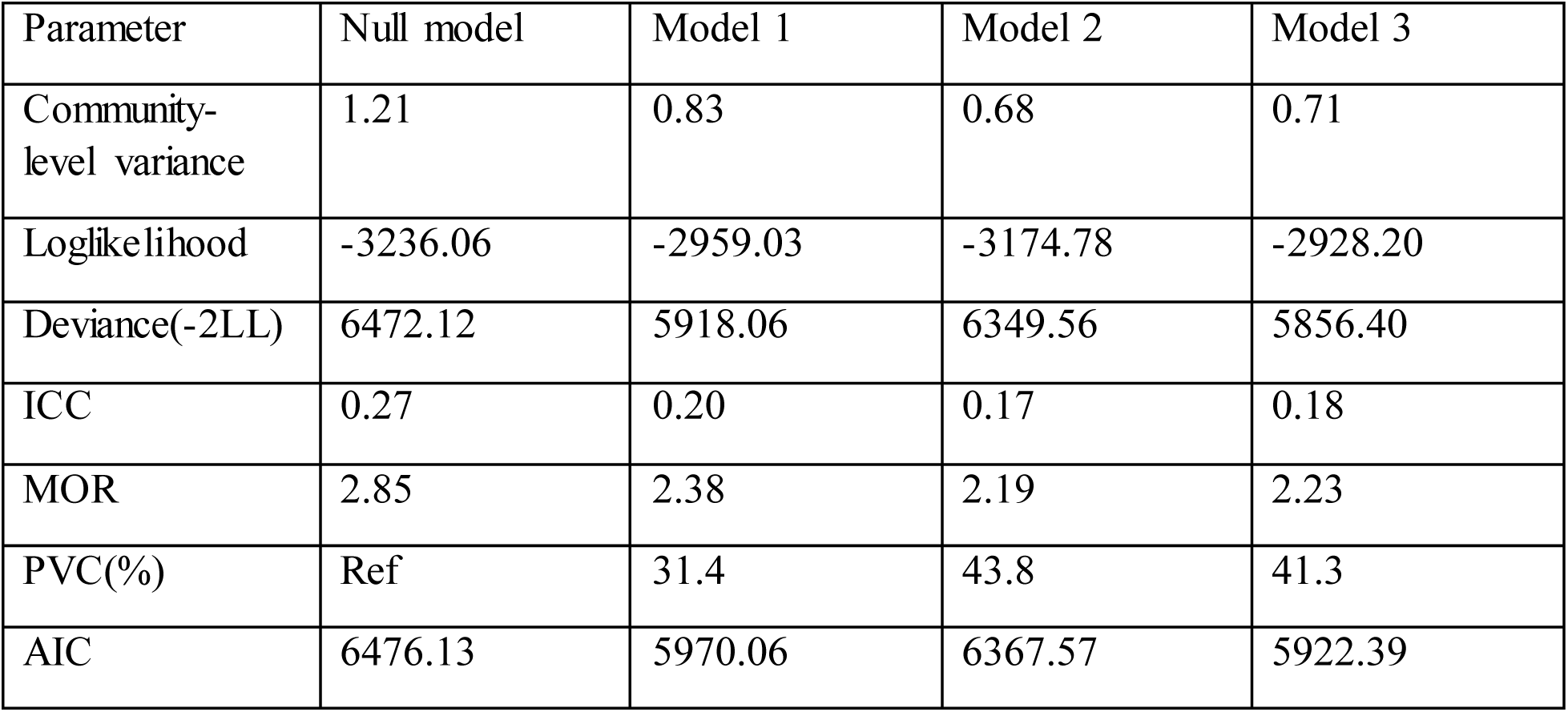
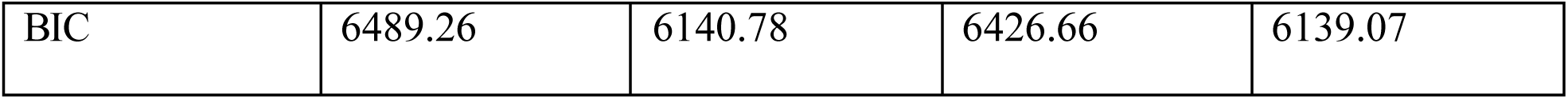
Random effect Analysis Result.

| Parameter | Null model | Model 1 | Model 2 | Model 3 |
| --- | --- | --- | --- | --- |
| Community-level variance | 1.21 | 0.83 | 0.68 | 0.71 |
| Loglikelihood | -3236.06 | -2959.03 | -3174.78 | -2928.20 |
| Deviance(-2LL) | 6472.12 | 5918.06 | 6349.56 | 5856.40 |
| ICC | 0.27 | 0.20 | 0.17 | 0.18 |
| MOR | 2.85 | 2.38 | 2.19 | 2.23 |
| PVC(%) | Ref | 31.4 | 43.8 | 41.3 |
| AIC | 6476.13 | 5970.06 | 6367.57 | 5922.39 |
| BIC | 6489.26 | 6140.78 | 6426.66 | 6139.07 |

### Random-effect Analysis Result

The null model showed that 27% total variability (ICC) for utilization of modern contraceptives was caused by variations among clusters, with the rest remaining unexplained due to within-cluster variability. In addition, 2.85(MOR) – the null model explains difference between clusters in modern contraceptives.

Finally, in multilevel model, PVC revealed that the full model explained 41.3% of the variability in modern contraception use. Model 3 was chosen as the best fit since it had the highest loglikelihood (lowest deviance) (Table 7).

**Model fit statistics**: the best model is the one with the lowest AIC/BIC and the highest log likelihood. Accordingly, model 3 is the best fitted model with smallest AIC and highest log likelihood.

The random effects analysis (multilevel binary logistic regression) demonstrated the contribution of community-level variation. The null models community-level variance was 1.21, which reduced to 0.71 in the full model. The ICC decreased from 0.27 to 0.18, indicating that 18% of the variance in modern contraceptive utilisation was attributable to community-level clustering after accounting for individual and community-level factors.

The proportional change in variance (PCV) showed that 41.3% of the community-level variation was explained in the full model. The median odds ratio (MOR) declined from 2.85 to 2.23, suggesting reduced disparity between communities.

## Discussion

In this study, we surveyed 5,250 reproductive-age women on their use of modern contraception and revealed a prevalence of 2188(41.7%). The magnitude of this study is consistent with findings from the 2019 Ethiopia Mini Demographic and Health Survey (EMDHS), which reported a modern contraceptive prevalence rate of 41% among married women of reproductive age (1). These consistent findings might be attributed to similarities in data collection tools, demographic characteristics, and national reproductive health strategies. The latest prevalence was higher than previous research in Ethiopia (28%)(11),20.4%(12), Gondar (41.2%)(13), Ghana (21%) (26), Central Africa(3.5%)(27).

However, the findings of this study were lower than those of studies conducted in, Boditi(28), Cameroon(29), Zimbabwe(30), Uganda(31), Comoros(32) and india(33). By using nationally representative PMA 2023 data and a multilevel statistical method, this study determined the magnitude and factors associated with modern contraceptive use among Ethiopian women of reproductive age. The result of this study reveal significant associations between modern contraception use and a variety of individual and community-level characteristics.

According to the findings of this study, older women aged from 35 to 49 years were less likely to use modern contraceptives [AOR = 0.50] as compared to those younger aged women(15 to 24 years), which aligns with studies from Ghana(34) and Ethiopia(18) that also noted decreased contraceptive use with increasing age. This may be explained by reduced fertility intentions or perceived reduced risk of pregnancy among older women. This finding is also consistent with research conducted in Sub-Saharan Africa, where older women, particularly those toward the end of their reproductive years, may believe they are less likely to become pregnant or have attained their preferred family size, reducing demand for contraception. On the other hand, for financial or educational reasons, younger women might be more inclined to postpone or space out pregnancies(35).

women who were not married had lower odds of utilizing modern contraceptives (AOR = 0.45) as compared to those married women, which aligns with studies conducted in Ethiopia(18) and Kenya(36), where married women were more likely to use modern contraceptives due to increased desire to space or limit births and better spousal communication. This aligns with evidence that married women are more likely to use contraception because they have better access to social support and family planning services (37). Particularly in conservative cultures, single women may face stigma or have less autonomy when seeking reproductive health care (3).

Women living in a moderate household size with 4 to 5 members had higher odds of utilizing modern contraceptives (AOR = 1.44) as compared to those women living in 1 to 3 household member. This may reflect greater awareness and discussion of family planning in larger households or increased motivation to limit further births due to economic pressures(38).

Women with more than 5 children were less likely to use modern contraceptives (AOR = 0.54) as compared to those with 0-2 children. This is consistent with studies showing that high-parity women may believe they have completed their desired family size or may rely on traditional methods (39). However, this also raises concerns about unmet need for contraception among multiparous women.

Women who had mixed feelings about getting pregnant had 2.56 times more odds utilizing modern contraceptives (AOR = 2.56) as compared to those women feel happy. Women who would be unhappy if pregnant had 2.13 times more odds of using modern contraceptives (AOR = 2.13) as compared to those women feel happy. Those who felt unhappy or mixed about a pregnancy had significantly higher odds of contraceptive use compared to those who would be happy. This underscores the role of pregnancy intentions in contraceptive behavior, as seen in other studies where ambivalence or negative feelings toward pregnancy drive contraceptive uptake (3, 67).

Women who visited health facility had higher odds of utilizing modern contraceptives (AOR = 1.53) as compared to those women who did not. Health facility visits increased the odds of contraceptive use by 53%, reinforcing the importance of healthcare access in promoting family planning. This finding aligns with studies conducted in Senegal(41) and Sierra Leone(42). Facility-based interventions, including counseling and service provision, have been shown to improve contraceptive adoption (43).

Women with moderate FP knowledge had higher odds of using modern contraceptives (AOR = 1.35) as compared to those women with poor knowledge. Family planning knowledge also played a role, with women having moderate knowledge showing higher contraceptive use than those with poor knowledge. This aligns with evidence that even basic awareness can improve contraceptive uptake, though comprehensive education yields better outcomes (44).

Women in the pastoralist regions were less likely to use modern contraceptives (AOR = 0.31) as compared to those women in the agrarian regions. Regional differences were evident, with women in pastoralist regions being 69% less likely to use contraceptives compared to agrarian regions. This may reflect disparities in healthcare access, cultural norms, or economic priorities, as pastoralist communities often have limited health infrastructure and higher mobility, reducing service utilization (45).

Interestingly, the study found that Muslim women were more likely to use modern contraceptives (AOR = 1.48) as compared to christians women. This contradicts previous studies that reported decreased contraceptive use among Muslim women(46). The disparity could be due to regional differences in behavior, interpretations of religious teachings, and targeted family planning initiatives.

Women who resides in communities with high level of women’s education had increased odds of using modern contraceptives (AOR = 1.82) as compared to those women in low education communities. This supports existing evidence that female education enhances reproductive autonomy and health-seeking behavior (47).

Women who reside in communities with high media exposure had increased odds of using modern contraceptives (AOR = 1.42) as compared to those women in low media exposure communities. It is consistent with findings that mass media campaigns effectively disseminate family planning information (48).

The multilevel model explained 41.3% of the variance in contraceptive use at the community level, with the intraclass correlation coefficient (ICC) decreasing from 27% to 18% after accounting for relevant factors. This shows that factors at both the individual and community levels have a significant impact on contraceptive behavior, underlining the need for multifaceted interventions.

This study has several strengths and limitations. Major strength of this study is the use of a recent, high-quality, and nationally representative Performance Monitoring for Action (PMA) Ethiopia 2023 data, which increases the generalizability of findings. Robust weighted sample size (n = 5,250) increases statistical power and the reliability of the results. Furthermore, the application of multilevel analysis in this study allows for the assessment of both individual– and community-level factors. However, the study also has limitations. Cross-sectional design restricts the study’s ability to establish causal relationships. The reliance on self-reported data may introduce recall bias, and unmeasured confounders such as the respondent’s occupation were not included, which could affect modern contraceptive utilization.

## Conclusion

Modern contraceptive utilization in Ethiopia remains suboptimal, which is below national targets. Utilization of modern contraceptives among reproductive-age females in Ethiopia influenced by both individual and community-level factors. Household member size, reaction if got pregnant now, visited facility, family planning knowledge, religion, community women education and media exposure were positively associated predictors. However, age, marital status, parity and region were negatively associated, which is less likely to use modern contraceptives.

### Implication of the Study

Modern contraceptive utilization is a key public health priority in Ethiopia, as it plays a critical role in improving maternal and child health outcomes and supporting women’s reproductive rights. The prevalence of modern contraceptive use among Ethiopian women of reproductive age (15-49 years) remains relatively low, and unmet needs for contraception persist across various regions and subpopulations(49).

Previous studies conducted in Ethiopia have primarily focused on individual-level factors using older datasets, such as the EDHS 2019 surveys. Existing studies, predominantly based on the Ethiopian Demographic and Health Survey (EDHS), often highlight limitations due to the unavailability of essential variable. However, these findings may not fully reflect the current realities, especially considering changes in service delivery models, community awareness, and family planning policies.

By using a recent PMA Ethiopia(2023) data set, variables missed in the previous researches will be considered. Additionally, limited attention has been given to community-level determinants, such as community family planning knowledge, which can shape social norms and access to services. These factors are crucial for understanding the determinants of contraceptive use.

This study, therefore, seeks to leverage the strengths of the PMA Ethiopia dataset to generate nuanced insights into modern contraceptive utilization and its associated factors among reproductive-age women, addressing gaps in existing literature and informing targeted, evidence-based interventions.

## Declarations

### Ethics approval

Ethical approval was not required as the study used publicly available, de-identified secondary data obtained from the PMA data repository. The PMA Ethiopia survey was conducted strictly under the ethical rules and regulations of world health organization and IRB of Ethiopian Public Health Institution (EPHI). Informed consent was obtained from respondents during the data collection process of PMA Ethiopia on data collection on November 2023. PMA survey has been also conducted after obtained ethical approval from Bloomberg School of Public Health at Johns Hopkins University in Baltimore, USA.

## Consent for publication

N/A not applicable

## Availability of data and materials

The data are freely available for public on Johns Hopkins University (JHU) Data Repository, which is open-access platform.

## Competing interests

the authors declare that they have no competing interest.

## Funding

The authors did not obtain any funding.

## Authors contribution

AD obtained the data; conducted the data curation and the analysis; draft the original manuscript and wrote the final draft, interpreted the results and critically revised the manuscript. SAD contributed to conceptualization of the study and interpretation of the results along with critically reviewing the final manuscript. MA contributed to critically reviewing the final manuscript version. All authors reviewed and approved the final manuscript.

## Data Availability

The data is publicly available on Johnhopkins university data repository.

https://archive.data.jhu.edu/

## List of Abbreviations

AIC: Akaike Information Criteria
AOR: Adjusted Odds Ratio
BIC: Bayesian Information Criteria
CI: Confidence Interval
CPR: Contraceptive Prevalence Rate
CS: Cross-sectional
EA: Enumeration Areas
EDHS: Ethiopian Demographic Health Survey
EHNRI: Ethiopian Health and Nutrition Research Institute
FP: Family Planning
HH: House Hold
ICC: Interclass Correlation
IRB: Institutional Review Board
IUD: Intra Uterine Device
MOR: Median Odds Ratio
NGO: Non-Governmental Organization
PMA: Performance Monitoring Action
PVC: Proportional Change Variance
SDG: Sustainable Development Goal
SRH: Sexual and Reproductive Health
SSA: Sub-Saharan Africa
SSNP: South Nations, Nationality and People region
STATA: Data Analysis and Statistical Software
VIF: Variance Inflation Factors
WHO: World Health Organization

## Acknowledgement

We have gotten the data from Johns Hopkins University(JHU) data repository. So, our deepest gratitude go to Johns Hopkins University for providing us such valuable data in open-access platform. Also, we would like to express our heartfelt gratitude to PMA Ethiopia team and respondents of PMA Ethiopia(2023) survey.

## Notes

### Competing Interest Statement

The authors have declared no competing interest.

### Clinical Protocols

https://archive.data.jhu.edu/

### Funding Statement

No funding for the study.

## REFERENCES

1. Ethiopian Public Health Institute (EPHI), ICF. Ethiopia Mini Demographic and Health Survey 2019: Final Report [Internet]. 2021. 1–207 p. Available from: https://dhsprogram.com/pubs/pdf/FR363/FR363.pdf

2. World Health Organization (WHO). World Family Planning [Internet]. United Nations. 2022. 43 p. Available from: https://www.un.org/en/development/desa/population/publications/pdf/family/WFP2017_Highlights.pdf

3. Sedgh G, Hussain R. Reasons for Contraceptive Nonuse among Women Having Unmet Need for Contraception in Developing Countries. Stud Fam Plann. 2014;45(2):151–69.

4. Negash BT, Chekol AT, Wale MA. Modern contraceptive method utilization and determinant factors among women in Ethiopia: Multinomial logistic regression mini-EDHS-2019 analysis. Contracept Reprod Med. 2023;8(1):1–9.

5. Health S. Sexual and Reproductive Health. J Obstet Gynaecol Res. 2023;49:138–44.

6. UN. World Family Planning. United Nations. 2022. 43 p.

7. Afriyie P, Tarkang EE. Factors influencing use of modern contraception among married women in Ho west district, Ghana: Descriptive cross-sectional study. Pan Afr Med J. 2019;33:1–11.

8. Njotang PN, Yakum MN, Ajong AB, Essi MJ, Akoh EW, Mesumbe NE, et al. Determinants of modern contraceptive practice in Yaoundé-Cameroon: A community based cross sectional study. BMC Res Notes. 2017;10(1):4–9.

9. United Nation. The 2030 Agenda and the Sustainable Development Goals An opportunity for Latin America and the Caribbean Thank you for your interest in this ECLAC publication [Internet]. 2018. 1–94 p. Available from: https://repositorio.cepal.org/bitstream/handle/11362/40156/25/S1801140_en.pdf

10. Worku AG, Tessema GA, Zeleke AA. Trends of modern contraceptive use among young married women based on the 2000, 2005, and 2011 Ethiopian demographic and health surveys: A multivariate decomposition analysis. PLoS One. 2015;10(1):1–14.

11. Hailegebreal S, Dileba Kale T, Gilano G, Haile Y, Endale Simegn A. Modern contraceptive use and associated factors among reproductive-age women in Ethiopia: multilevel analysis evidence from 2019 Ethiopia mini demographic and health survey. J Matern Neonatal Med [Internet]. 2023;36(2). Available from: 10.1080/14767058.2023.2234067

12. Mamo, Nigatu, Gebre, Zerihun, Kura E. Modern contraceptive utilization and associated factors among reproductive-age women in Ethiopia: evidence from 2016 Ethiopia demographic and health survey. BMC Womens Health. 2020;

13. Oumer M, Manaye A, Mengistu Z. Modern Contraceptive Method Utilization and Associated Factors Among Women of Reproductive Age in Gondar City, Northwest Ethiopia. Open Access J Contracept. 2020;Volume 11:53–67.

14. Tesema ZT, Tesema GA, Boke MM, Akalu TY. Determinants of modern contraceptive utilization among married women in sub-Saharan Africa: multilevel analysis using recent demographic and health survey. BMC Womens Health [Internet]. 2022;22(1):1–11. Available from: 10.1186/s12905-022-01769-z

15. Mankelkl G, Kassaw AB, Kinfe B. Factors associated with modern contraceptive utilization among reproductive age women in Kenya; evidenced by the 2022 Kenyan demographic and health survey. Contracept Reprod Med. 2024;9(1):1–8.

16. Beyene KM, Bekele SA, Abu MK. Factors affecting utilization of modern contraceptive methods among women of reproductive age in Ethiopia. PLoS One [Internet]. 2023;18(11 November):1–12. Available from: 10.1371/journal.pone.0294444

17. Apanga PA, Kumbeni MT, Ayamga EA, Ulanja MB, Akparibo R. Prevalence and factors associated with modern contraceptive use among women of reproductive age in 20 African countries: A large population-based study. BMJ Open. 2020;10(9):1–12.

18. Zeleke GT, Zemedu TG. Modern contraception utilization and associated factors among all women aged 15–49 in Ethiopia: evidence from the 2019 Ethiopian Mini Demographic and Health Survey. BMC Womens Health [Internet]. 2023;23(1):1–7. Available from: 10.1186/s12905-023-02203-8

19. Martin V, Msuya SE, Kapologwe N, Damian DJ, John B, Mahande MJ. Prevalence and Determinants of Modern Contraceptive Methods Use among Women of Reproductive Age (15 – 49 Years) in Rural Setting: A Case of Kishapu District, Shinyanga Region. Adv Sex Med. 2019;09(04):53–66.

20. Abiye AA, Fekede B, Jemberie AM, Molla BA, Tolla BK, Tefera BS, et al. Modern Contraceptive Use and Associated Factors among Reproductive Age Group Women in three Peri-Urban Communities in Central Ethiopia. J Drug Deliv Ther. 2019;9(6-s):93–102.

21. Ahinkorah BO, Budu E, Aboagye RG, Agbaglo E, Arthur-Holmes F, Adu C, et al. Factors associated with modern contraceptive use among women with no fertility intention in sub-Saharan Africa: evidence from cross-sectional surveys of 29 countries. Contracept Reprod Med. 2021;6(1):1–13.

22. Abdu Yesuf K. Modern contraceptive utilization and associated factors among younger and older married youth women in Ethiopia: Evidence from Ethiopia Mini Demographic and Health Survey 2019. PLoS One [Internet]. 2024;19(5):e0300151. Available from: 10.1371/journal.pone.0300151

23. Debebe S, Limenih MA, Biadgo B. Modern contraceptive methods utilization and associated factors among reproductive aged women in rural Dembia District, northwest Ethiopia: Community based cross-sectional study. Int J Reprod Biomed. 2017;15(6):367–74.

24. Shiferaw S, Principal MPH. Study Title – Performance Monitoring for Action Ethiopia (Pma Ethiopia). 2023;(October).

25. Addis Ababa University School of Public Health; and the Williams H. Gates Sr. Institute for Population and Reproductive Health at the Johns Hopkins Bloomberg School of Public Health. Performance Monitoring for Action Ethiopia (PMA-ET) 2023 Cross-sectional Household and Female Survey (Version 2.0), PMAET-2023CS-HQFQ. 2023.

26. Tegegne TK, Chojenta C, Forder PM, Getachew T, Smith R, Loxton D. Spatial variations and associated factors of modern contraceptive use in Ethiopia: a spatial and multilevel analysis. BMJ Open. 2020;10(10):1–11.

27. Boadu I. Coverage and determinants of modern contraceptive use in sub-Saharan Africa: further analysis of demographic and health surveys. Reprod Health [Internet]. 2022;19(1):1–11. Available from: 10.1186/s12978-022-01332-x

28. Gebremeskel F. Prevalence of Modern Contraceptive Utilization and Associated Factors Among Women of Reproductive Age Group at Boditi Town, Wolayita Zone, SNNPR, Ethiopia. Am J Nurs Sci. 2017;6(6):447.

29. Tchakounté Tchuimi D, Kamga BF. The effect of women’s bargaining power within couples on contraceptive use in Cameroon. Gates Open Res. 2024;4:1–23.

30. Joseph, Lasong., Bassoumah, Bougangue., Yaa, Nyarko A. Modern contraceptive use among women of reproductive age in Zimbabwe: analysis of 1999-2015 Zimbabwe Demographic Health Survey. Eur J Contracept Reprod Heal Care [Internet]. 2022; Available from: https://pubmed.ncbi.nlm.nih.gov/35959761/

31. Towongo MF, Kelepile M. Prevalence, distribution and factors associated with modern contraceptive use among women of reproductive age in Uganda: evidence from UDHS 2016. Contracept Reprod Med. 2024;9(1):1–17.

32. Tessema ZT, Teshale AB, Tesema GA, Yeshaw Y, Worku MG. Pooled prevalence and determinants of modern contraceptive utilization in East Africa: A Multi-country Analysis of recent Demographic and Health Surveys. PLoS One [Internet]. 2021;16(3 March):1–16. Available from: 10.1371/journal.pone.0247992

33. Agrawal R, Mishra M, Rehman T, Surendran G, Sinha A, Kanungo S, et al. Utilization of modern temporary contraceptive methods and its predictors among reproductive-aged women in India: insights from NFHS-5 (2019–21). Front Glob Women’s Heal. 2023;4(October):1–11.

34. Takyi A, Sato M, Adjabeng M, Smith C. Factors that influence modern contraceptive use among women aged 35 to 49 years and their male partners in Gomoa West District, Ghana: a qualitative study. Trop Med Health [Internet]. 2023;51(1). Available from: 10.1186/s41182-023-00531-x

35. Bongaarts, J., & Hardee K. The Role of Public-Sector Family Planning Programs in Meeting the Demand for Contraception in Sub-Saharan Africa. Int Perspect Sex Reprod Heal [Internet]. 2017;43(2),:41–50. Available from: 10.1363/43e3917

36. Ochako R, Temmerman M, Mbondo M, Askew I. Determinants of modern contraceptive use among sexually active men in Kenya. Reprod Health. 2017;14(1):1–15.

37. Wulifan JK, Brenner S, Jahn A, De Allegri M. A scoping review on determinants of unmet need for family planning among women of reproductive age in low and middle income countries. BMC Womens Health [Internet]. 2016;16(1). Available from: 10.1186/s12905-015-0281-3

38. Eliason S, Awoonor-Williams JK, Eliason C, Novignon J, Nonvignon J, Aikins M. Determinants of modern family planning use among women of reproductive age in the Nkwanta district of Ghana: A case-control study. Reprod Health. 2014;11(1):1–10.

39. Ahinkorah BO, Hagan JE, Seidu AA, Sambah F, Adoboi F, Schack T, et al. Female adolescents’ reproductive health decision-making capacity and contraceptive use in sub-Saharan Africa: What does the future hold? PLoS One [Internet]. 2020;15(7 July):1–20. Available from: 10.1371/journal.pone.0235601

40. Nancy L Stanwood, Susan E Cohn, Jennifer R. Heiser, MaryAnn Pugliese. Contraception and fertility plans in a cohort of HIV-positive women in care. Contraception. 2007;75(4):294–298.

41. Endale HT, Negash HK, Tesfaye W, Hasen FS, Asefa T, Gelaw DT, et al. Utilization of modern contraceptive methods among women of reproductive age in Senegal: A multilevel mixed-effects analysis. PLoS One [Internet]. 2025;20(5 May):35–44. Available from: 10.1371/journal.pone.0323899

42. Sserwanja Q, Turimumahoro P, Nuwabaine L, Kamara K, Musaba MW. Association between exposure to family planning messages on different mass media channels and the utilization of modern contraceptives among young women in Sierra Leone: insights from the 2019 Sierra Leone Demographic Health Survey. BMC Womens Health [Internet]. 2022;22(1):1–10. Available from: 10.1186/s12905-022-01974-w

43. Prata N, Fraser A, Huchko MJ, Gipson JD, Withers M, Lewis S, et al. Women’s Empowerment and Family planning. J Biosoc Sci. 2017;49(6):713–43.

44. Do M, Kurimoto N. Women’s empowerment and choice of contraceptive methods in selected African countries. Int Perspect Sex Reprod Health. 2012;38(1):23–33.

45. Mekonnen W, Worku A. Determinants of low family planning use and high unmet need in Butajira District, South Central Ethiopia. Reprod Health [Internet]. 2011;8(1):37. Available from: http://www.reproductive-health-journal.com/content/8/1/37

46. Gebrekidan H, Alemayehu M, Debelew GT. Individual and community level factors associated with modern contraceptive utilization among women in Ethiopia: Multilevel modeling analysis. PLoS One [Internet]. 2024;19(5):1–20. Available from: 10.1371/journal.pone.0303803

47. Chola L, McGee S, Tugendhaft A, Buchmann E, Hofman K. Scaling up family planning to reduce maternal and child mortality: The potential costs and benefits of modern contraceptive use in South Africa. PLoS One. 2015;10(6):1–16.

48. Hutchinson PL, Meekers D. Estimating Causal Effects from Family Planning Health Communication Campaigns Using Panel Data: The “Your Health, Your Wealth” Campaign in Egypt. PLoS One. 2012;7(9).

49. Westoff CF. Unmet need for modern contraceptive methods. DHS Anal Stud No 28 [Internet]. 2012;(September). Available from: http://dhsprogram.com/pubs/pdf/AS28/AS28.pdf

